# Coaching visits and supportive supervision for primary care facilities to improve malaria service data quality in Ghana: an intervention case study

**DOI:** 10.1101/2024.08.07.24311636

**Authors:** Amos Asiedu, Rachel A. Haws, Wahjib Mohammed, Joseph Boye-Doe, Charles Agblanya, Raphael Ntumy, Keziah Malm, Paul Boateng, Gladys Tetteh, Lolade Oseni

## Abstract

Effective decision-making for malaria prevention and control depends on timely, accurate, and appropriately analyzed and interpreted data. Poor quality data reported into national health management information systems (HMIS) prevent managers at the district level from planning effectively for malaria in Ghana. We analyzed reports from data coaching visits and follow-up supervision conducted to 231 health facilities in six of Ghana’s 16 regions between February and November 2021. The visits targeted health workers’ knowledge and skills in malaria data recording, HMIS reporting, and how managers visualized and used HMIS data for planning and decision making. A before-after design was used to assess how data coaching visits affected data documentation practices and compliance with standards of practice, quality and completeness of national HMIS data, and use of facility-based malaria indicator wall charts for decision-making at health facilities. The percentage of health workers demonstrating good understanding of standards of practice in documentation, reporting and data use increased from 72 to 83% (p<0.05). At first follow-up, reliability of HMIS data entry increased from 29 to 65% (p<0.001); precision increased from 48 to 78% (p<0.001); and timeliness of reporting increased from 67 to 88% (p<0.001). HMIS data showed statistically significant improvement in data completeness (from 62 to 87% (p<0.001)) and decreased error rate (from 37 to 18% (p<0.001)) from baseline to post-intervention. By the second follow-up visit, 98% of facilities had a functional data management system (a 26-percentage-point increase from the first follow-up visit, p<0.0001), 77% of facilities displayed wall charts, and 63% reported using data for decision-making and local planning. There are few documented examples of data coaching to improve malaria surveillance and service data quality. Data coaching provides support and mentorship to improve data quality, visualization, and use, modeling how other malaria programs can use HMIS data effectively at the local level.

## Introduction

Efficient and sustainable decision-making for disease prevention and control depends on timely, accurate, well-collected, appropriately analyzed, and effectively interpreted data (1). In low- and middle-income countries (LMICs), routine data for decision-making are available primarily through each country’s national Health Management Information System (HMIS) (2). It is important that data collected are of sufficient quality to allow regular tracking of progress for monitoring impact, scaling up interventions, and strengthening health systems (3–6). Routine service data are often poor quality or incomplete, and may be misleading or of limited use for planning and decision-making (3,7). Concerns about data quality often limit the use of routine service data for prioritizing or targeting program activities, as stakeholders do not trust the integrity of aggregated datasets enough to rely on their analysis for decision-making (7,8). Recent efforts by ministries of health and stakeholders to improve the quality of HMIS data have prompted interventions at the district level to improve data quality and use, including technological innovations, capacity strengthening, data quality audits, and supportive supervision approaches (9–11).

Gaps in data quality illustrate the need for improved quality of data to inform decision making for malaria prevention, control, and elimination efforts (7). An assessment of National Malaria Strategic Plans (NMSPs) in 22 countries in sub-Saharan Africa showed that challenges in surveillance, monitoring, and evaluation (SME) included weak health information systems, incomplete data collection, poor data quality, inadequate data management and use and a lack of integration of monitoring systems (12). Data management issues such as failing to report certain malaria indicators and data reporting delays generally remained unaddressed in subsequent NMSPs. Poor motivation, inadequate supervision, absent feedback mechanisms, and competing disease-specific reporting requests that burden staff contribute to low quality in routine data reporting (13).

Ensuring that high-quality routine data are collected on malaria service provision will be essential to meet aggressive targets set by the World Health Organization (WHO) and RBM Partnership to End Malaria to move countries from malaria control to pre-elimination by 2030 and reduce cases by 90 percent (14,15). Ghana aims to move six districts from malaria control to pre-elimination by 2025. Studies from countries that have reached the malaria elimination stage have proposed that data-driven decision-making and improved surveillance are essential to address technical, operational, and financial challenges associated with pursuing the goal of elimination, with surveillance as a core intervention (16). Surveillance is generally weakest in countries with a high malaria burden, underlining the importance of the need for a robust surveillance system (14). It is essential that quality data on malaria service delivery are available and accessible to policy makers, facility managers, and health workers (17).

In Ghana, routine data at the facility level are reported monthly in the national electronic HMIS using open-source District Health Information System version 2 (DHIS2) software, which was introduced in 2012 to replace paper-based records and includes data validation, visualization and analysis tools. In Ghana, the DHIS2-based platform is called District Health Information Management System version 2 (DHIMS2). Since DHIMS2 became available, the percentage of health facilities submitting required monthly reports has risen from 11% in 2012 to 91.6% in 2021 (18). Health workers record provision of malaria-related services using specific recording and reporting tools in DHIMS2. Regular review of DHIMS2 data can be used to assess health system performance and for local planning and decision-making.

Supportive supervision visits, regional holistic assessments, and SME Technical Working Group meetings have revealed that health workers in Ghana, especially those posted at primary care facilities, lack understanding of the Ghana Health Service standards of practice regarding health information management and how to record some data elements in registers and reporting forms. Additionally, reviews of routine DHIMS2 data have revealed disparities between and within regions in terms of data quality. This problem has led to errors in data capture, extraction, and reporting. As has been documented in other countries, health workers generally underutilize available data visualization tools and techniques to facilitate the use of local facility data for planning and decision making (7,19–21).

Primary health care workers at Community Health Planning and Services (CHPS) compounds and health centers in Ghana play a critical role in collecting, documenting, and reporting malaria service data. Service providers enter patient-level information in facility registers and collate data into monthly reporting forms; they also complete monitoring charts to guide clinical meetings and procurement of supplies. Data managers at district level and in some larger and higher-volume facilities conduct data verification and validation of monthly reporting forms and registers with service providers, enter data into DHIMS2, and review data for facility performance and local planning. In facilities without data managers, service providers often perform these roles. Coaching them in proper data documentation could improve the quality of the data they collect, reducing the risk of errors, missing information, or duplicate entries (22). Strengthening capacity in good data management practices can ensure that routine data is reliable and usable for monitoring progress, evaluating programs, and informing best practices. Effective data management can also streamline processes and improve efficiency, freeing up time for health care providers to focus on patient care. Training health workers in basic data visualization techniques can empower them to track progress and make data-driven decisions at the local level. The U.S. President’s Malaria Initiative (PMI) Impact Malaria project conceptualized data coaching visits as a means to improve malaria data quality and data use through targeted support to primary facilities with low baseline levels of data quality and completeness.

This case study evaluates how data coaching visits to targeted health facilities affected health workers’ knowledge and skills in malaria data recording, extraction and reporting to DHIMS2, and subsequent changes in how managers visualized and used DHIMS2 data for planning and decision making.

## Data coaching intervention and evaluation methods

We used a before-after design to assess the effect of data coaching visits on data documentation practices and compliance with standards of practice for DHIMS2, quality and completeness in DHIMS2, and use of facility-based malaria indicator wall charts for decision-making at 231 health facilities in six of Ghana’s 16 regions. Follow-up visits and analysis of key malaria indicators in DHIMS2 were used to assess longer-term changes in data documentation, reporting, and use for decision-making. Findings from this case study illustrate how health workers’ skills in malaria data documentation, reporting, and validation can be built and sustained.

Data coaching visits to facilities were designed to strengthen data managers’ and service providers’ competency in proper documentation in registers, accurate transfer of data onto reporting forms and into DHIMS2, and ability to use tools that track and present data on malaria indicators. Using the Plan-Do-Study-Act (PDSA) continuous improvement cycle (23), which is an iterative quality improvement approach that identifies problems and develops and tests potential solutions—“change ideas”—to these problems, managers were guided during these visits to develop change ideas to address identified data gaps in their districts and facilities (Fig 1).

**Fig 1.**
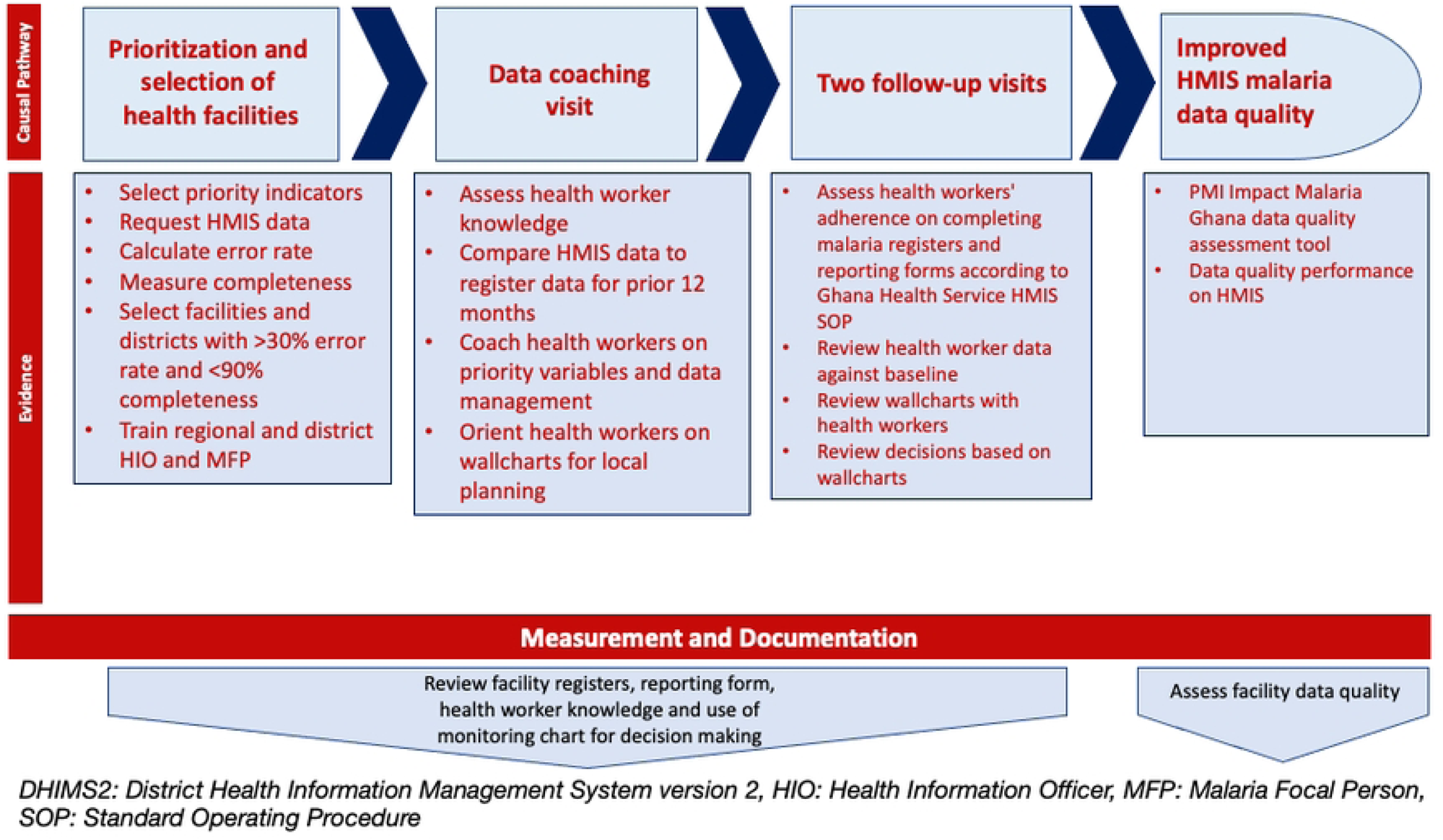
Implementation framework.

Following the WHO High Burden High Impact (HBHI) strategy, we prioritized primary health facilities—health centers and CHPS—with poor data quality to receive data coaching visits. The intervention prioritized these facilities by using a prioritization matrix to select facilities with error rates exceeding 30 percent and data completeness rates less than 90 percent on three malaria indicators in national DHIMS2 data from August 2020 to January 2021. Error rate was calculated as the number of indicators flagged with errors during the data validation exercise divided by the total number of validated indicators. Completeness rate was calculated as the number of data elements reported in DHIMS2 divided by the total number of expected data elements. The three indicators were: 1) number of suspected malaria cases at outpatient department (OPD) tested for malaria using rapid diagnostic tests (RDT) or microscopy; 2) number of confirmed malaria cases, disaggregated by test type: RDT and microscopy; and 3) confirmed uncomplicated malaria cases treated with antimalarial artemisinin-combination therapy (ACT). A total of 231 out of 605 facilities in 52 districts in 6 of Ghana’s 16 regions were prioritized using the matrix.

Prior to the coaching visits, PMI Impact Malaria staff, in collaboration with National Malaria Elimination Program and Policy, Planning, Monitoring & Evaluation Division (PPMED) staff from the Ghana Ministry of Health, trained regional and district level malaria focal persons and health information officers on the data coaching tools and DHIMS2 standards of practice. Two teams made up of three staff (the district and regional health information officer and a malaria focal person) then provided data coaching visits to all facilities, with each team visiting two facilities per day and spending an average of 4 hours in each facility.

During each coaching visit, health workers were given a pre-test on knowledge about the various malaria-related registers and reporting forms, variables in registers (e.g., definition of old versus new cases), tools for data validation, and use of facility-based wall charts for planning. First, coaching teams reviewed facility data with health workers to compare the source (register) to the data reported to DHIMS2. Next, they coached health workers on register variables, completion of reporting forms, and data validation following DHIMS2 standards of practice. Coaching teams then presented a facility-based wall chart to explain proper documentation, reporting and the use of wall charts for local planning, demonstrating for participants how to complete and use malaria indicator wall charts to track the performance of malaria indicators at the facility level (See S1 Fig). During these visits, coaching teams also guided managers to develop change ideas using the PDSA continuous improvement cycle to address identified data gaps in their districts and facilities.

Coaching teams conducted two post-coaching follow-up visits three months and nine months after the coaching visit using the same assessment tools to assess improvement in data reporting, facility display of updated wall charts, and reported use of the dashboard and wall chart for decision-making.

To evaluate the intervention, we used a before-after design that assessed how the coaching visits improved health workers’ knowledge about malaria data variables in registers, their reporting practices, and their competence in visualizing and using data for local planning and decision making. We also evaluated health workers’ use of data for local planning over the course of the program and follow-up, and evaluated the experiences and perceptions of health workers and regional and district level supervisors who had participated in the data coaching activities via a satisfaction survey.

### Data collection

#### Facility performance monitoring system and quality of data being reported in DHIMS2

On three occasions—during the coaching visit (February 2021), the first follow-up visit (May 2021), and the second follow-up visit (November 2021)—we assessed whether facilities had functional data management systems conforming to GHS standards of practice, and health worker understanding of the GHS standard performance monitoring system and data quality. The assessment evaluated health worker understanding of the facility data management system and compliance to GHS standards of practice for documenting data in DHIMS2. Regional and district level health information officers conducted the assessments using a standardized USAID Data Quality Assurance tool programmed on the KoboCollect App (24) and stored all collected data in the PMI Impact Malaria database. The tool assessed data quality across five dimensions: validity, reliability, integrity, precision, and timeliness. To measure improvement in the performance monitoring system of the facilities, we compared assessment scores from the second follow-up visit to the first follow-up and the initial coaching visit.

#### Facility reporting data completeness and error rate in national DHIMS2 data

We also assessed how improvements in the facility performance monitoring system translated to changes in data reporting completeness and error rates in national DHIMS2 data after the coaching visits by comparing seven tracked national malaria case management indicators from baseline (0-6 months prior to the coaching visit) to data from monthly reporting forms and registers from each facility from three periods after the coaching visits: midline (1-9 months post-coaching visit), endline (10-12 months post-coaching visit), and post-endline (13-16 months post-coaching visit). These indicators included suspected cases of uncomplicated malaria, suspected cases of uncomplicated malaria tested, confirmed malaria cases, outpatients treated with antimalarials, outpatients treated with artemisinin-combination treatments, suspected uncomplicated malaria cases tested using RDTs, and suspected uncomplicated malaria cases tested using microscopy. Facilities collated quantitative facility-level service data, reported monthly to DHIMS2, for the 231 health facilities targeted for the coaching visit.

#### Data use for decision making and local planning

We assessed whether and how facilities visualized data for decision making and data use for planning at the coaching visit, and again at the first and second follow-up visits. We used the KoboCollect App to collect observations on whether wall charts were displayed, updated, and being used for decision-making about procurement of supplies, workflow, adherence to guidelines, in-service trainings, and health education priorities. All data were stored in the PMI Impact Malaria database. We measured improvement in the accurate updating of facility-based wall chart and data use for local planning (e.g., ITN distribution, reducing delays in service provision, and identifying staff training needs) by comparing assessment scores from the first follow-up visit to the coaching visit, and the second follow-up visit to the first.

#### Data coaching satisfaction survey

We conducted an after-action review from March-April 2022 using an online satisfaction survey. We sent a Google Forms link via WhatsApp to health workers at facilities that had received coaching visits (N=462), as well as health information officers (N=52) and malaria focal persons (N=52). The survey link was closed in March 2022 after a convenience sample of 250 participants had responded anonymously to the survey. Facilities were randomly selected, but inclusion criteria included smartphone ownership and internet access, which limited the sample by approximately 40%. The survey assessed the opinions, experience, and perspectives of health workers, health information officers, and malaria focal persons about how data coaching affected facilities’ performance systems, data quality, use of data for decision making and data reporting in DHIMS2.

### Data analysis

#### Facility performance monitoring system

We used a two-sample t-test at a 5% level of significance to detect differences in overall facility assessment scores between the first follow-up visit and the coaching visit, as well as between the second follow-up visit and the first follow-up visit.

#### Effect on service delivery

We used a paired t-test using Stata/IC version 14.2 to identify statistically significant changes over time in routine monthly DHIMS2 indicators on quality malaria case management (25). The analysis measured changes in data reporting completeness and error rates using the period 0-6 months prior to the coaching visit as a baseline, comparing this to indicators from midline, endline, and post-endline periods after the data coaching visit.

#### Data coaching satisfaction survey

The study team read through survey responses and highlighted emergent themes individually in Atlas.ti Version 7.5 using a thematic analysis approach (26). Themes and codes were reviewed and discussed throughout the analytic process, and refined or adapted as needed based on emergent information from the survey responses. Data were analyzed by type of participant, then discussed and summarized, noting differences between types of participants. The team then collated and reached consensus on themes through team discussions.

### Data management and confidentiality

No personally identifiable information was collected during the assessment. All secondary data included in the study were routine malaria service data that are publicly available. Project monitoring and evaluation data gathered during the coaching visits and follow-up visits included providers’ sex, cadre, data coaching performance assessment score, and evidence of data use for decision making. Names of providers and their facilities were not collected. The qualitative satisfaction survey was anonymous: names and facility identification of health workers were not collected as part of the survey. All data collected and retrieved for the study were stored securely in the password-protected PMI Impact Malaria database.

### Ethical considerations

This assessment obtained a non-human subjects research determination from the Johns Hopkins Bloomberg School of Public Health Institutional Review Board, Maryland, USA (IRB# 21543). Study participants who completed the satisfaction survey gave their written informed consent.

## Results

### Participating facilities, health workers, and data coaches

A total of 14 regional-level and 104 district-level supervisors, including malaria focal persons and health information officers from 52 districts across six regions, served as coaches and visited facilities. All 231 targeted health facilities received a coaching visit; 833 health workers (327 male; 506 female) participated in coaching visits at these facilities. Follow-up visits conducted 3 months and 9 months after coaching visits reached 1211 health workers at each visit (578 male; 633 female). The same number of health workers were reached at each follow-up visit, though the individuals reached at each follow-up visit varied. Twenty-three percent (N=54) of the 231 targeted facilities were health centers, and the rest (N=177) were CHPS.

### Facility performance monitoring system and data quality assessment

Almost all performance monitoring scores and DHIMS2 standard of practice compliance improved from baseline to first follow-up visit (Table 1). The percentage of health workers demonstrating good understanding of documenting, reporting and use increased from 72 to 83% (p<0.05). Sixty percent of facilities were using standard malaria registers at the initial coaching visit, but at the second follow-up visit, 95% of the facilities were using standard malaria registers. While the percentage of facilities with functional data storage and management systems only increased slightly between the coaching visit and first follow-up visit, 98% of facilities had a functional data storage and management system by the second follow-up visit, from 72% at first follow-up visit (p<0.0001).

**Table 1.** Changes in performance, adherence to standards of practice, and data quality after data coaching visit.

| Performance Monitoring System and Data Quality Assessment | Coaching visit score (N=833, %) | First follow-up visit score (N=1211, %) | Difference (First follow-up vs. coaching visit) (percentage points) | p-value | Second follow-up visit score (N=1211, %) | Difference (Second vs. first follow-up) (percentage points) | p-value |
| --- | --- | --- | --- | --- | --- | --- | --- |
| **Monitoring System** |  |  |  |  |  |  |  |
| Staff demonstrate good understanding of data recording, reporting and use | 72% | 83% | **11**<br>(2, 21) | 0.046 | 89% | 6<br>(-6, 18) | 0.353 |
| Functional facility data storage and management system | 68% | 72% | 4<br>(-7, 15) | 0.501 | 98% | **26**<br>(16, 36) | **<0.001\*** |
| **DHIMS2 Standards of Practice Compliance** |  |  |  |  |  |  |  |
| Availability of GHS DHIMS2 SOP | 36% | 48% | 12<br>(-0.4, 24) | 0.057 | 50% | 2<br>(-16, 20) | 0.825 |
| Staff able to differentiate between a new and old case* of malaria | 61% | 89% | **28**<br>(18, 37) | **<0.001\*** | 96% | 7<br>(-2, 15) | 0.169 |
| Staff report only 'New Cases' of malaria for the malaria section of OPD monthly reporting form | 83% | 94% | **11**<br>(4, 18) | **0.012\*** | 89% | -5<br>(-15, 5) | 0.301 |
| Staff report only 'Old Cases'* of malaria under repeat attendances in the OPD monthly reporting form | 73% | 88% | **15**<br>(6, 24) | **0.005\*** | 93% | 5<br>(-5, 15) | 0.364 |
| Reporting case management cascade (number suspected, tested with RDT or microscopy, confirmed positive, treated with ACT) in OPD monthly reporting form, with first three indicators disaggregated by pregnancy status | 80% | 93% | **13**<br>(5, 20) | **0.005\*** | 93% | 0<br>(-9, 9) | 1.000 |
| Errors in malaria data reported, have been tracked to their original source and mistakes corrected | 89% | 83% | 6<br>(-15, 3) | 0.166 | 89% | 6<br>(-6, 18) | 0.178 |
| Data is reviewed for accuracy before sending to the next level/entry into DHIMS2 | 42% | 65% | **23**<br>(11, 35) | **<0.001\*** | 82% | **17**<br>(2, 32) | **0.039\*** |
| Appropriate procedures used to ensure accurate data transferred from registers to reporting forms | 53% | 66% | **13**<br>(9, 25) | **0.041\*** | 89% | **23**<br>(10, 36) | **0.004\*** |
| Appropriate procedures for avoiding double counting of malaria data | 45% | 65% | **20**<br>(10, 32) | **0.002\*** | 80% | 15<br>(-1, 30) | 0.071 |
| Systems and procedures in place to ensure timely reporting of malaria data | 73% | 83% | 10<br>(-1, 20) | 0.068 | 89% | 6<br>(-6, 18) | 0.354 |

\* An "old case" is defined as a client who tested positive for malaria within 28 days of the first episode.

Compliance of health workers with almost all standards of practice for DHIMS2 reporting had improved at the first follow-up visit (Table 1). Improvements were statistically significant for four discrete skills: ability to differentiate between a new and old case of malaria, reporting only new cases on the malaria section of OPD morbidity, reporting only old cases under “repeat attendances” on OPD morbidity returns, and reporting case management cascades (number suspected, tested with RDT or microscopy, confirmed positive, treated with ACT) in OPD morbidity returns. Data quality also improved by the first follow-up visit, including increased reporting of errors and correcting mistakes, data reviewed for accuracy, procedures used to ensure accurate data from registers to reporting, and avoiding double counting of malaria data.

### Facility data quality reported to DHIMS2

Compared to baseline, the quality of data that facilities reported to DHIMS2 at the first follow-up visit improved significantly across three dimensions of quality (Table 2). Reliability increased from 29% to 65% (p<0.001); precision increased from 48% to 78% (p<0.001); and timeliness of reporting increased from 67% to 88% (p<0.001). By the second follow-up visit, reliability had increased from 65% to 89% (p<0.05). Integrity also improved from 66% at first follow-up visit to 90% at the second follow-up visit (p<0.05).

**Table 2.** Changes in USAID Data Quality Assurance tool scores on data quality dimensions after data coaching.

| Data Quality Assessment Dimension | Coaching visit score (N=833) | First follow-up visit* score (N=1211) | Percentage-point difference (first follow-up vs. coaching visit)<br>Mean (95% CI) | *p-value* | Second follow-up visit* score (N=1211) | Percentage-point difference (second vs. first follow-up)<br>Mean (95% CI) | *p-value* |
| --- | --- | --- | --- | --- | --- | --- | --- |
| Validity | 87% | 94% | +7<br>(-8, +14) | 0.079 | 94% | 0<br>(-11, +11) | 1.000 |
| Reliability | 29% | 65% | **+36**<br>**(+24, +48)** | **<0.000\*** | 89% | **+24**<br>**(+11, +37)** | **0.003\*** |
| Integrity | 54% | 66% | +12<br>(-2, +24) | 0.059 | 90% | **+24**<br>**(+11, +37)** | **0.002\*** |
| Precision | 48% | 78% | **+30%**<br>**(+19, +41)** | **<0.000\*** | 89% | +11<br>(-2, +23) | 0.116 |
| Timeliness | 67% | 88% | **+ 21%**<br>**(+11, +31)** | **<0.000\*** | 91% | +3<br>(-8, +14) | 0.597 |

First follow-up visit 3 months post-coaching; second follow-up visit 9 months post-coaching.

### Data completeness and error rate in DHIMS2

Data in DHIMS2 for the study facilities showed statistically significant improvement in data completeness, from 62% at baseline to 87% at midline (p<0.001) and decreased error rate, from 37% at baseline to 18% at midline (p<0.001). Error rates continued to drop in the period 10-12 months after the coaching visits compared to the period 1-9 months post-coaching visits (p<0.05). Completeness also improved, but the difference was not statistically significant.

**Table 3.** Error rates and completeness in DHIMS2 pre- and post-data coaching.

| **DHIMS2 Data Reporting** | **Baseline (0-6 mos pre-coaching)**<br>**n=138 600** | **Midline (1-9 mos post-coaching)**<br>**n=140 122** | **Percentage-point difference midline vs baseline** | **Endline (10-12 mos post-coaching)**<br>**n=139 433** | **Percentage-point difference endline vs midline** | **Post-endline (13-16 mos post-coaching)**<br>**n=139 433** | **Percentage-point difference post-endline vs endline** |
| --- | --- | --- | --- | --- | --- | --- | --- |
|  | Mean %<br>(+/- SD) | Mean %<br>(+/- SD) | Mean %<br>(95% CI)<br>*p-value* | Mean %<br>(+/-SD) | Mean %<br>(95% CI)<br>*p-value* | Mean %<br>(+/-SD) | Mean %<br>(95% CI)<br>*p-value* |
| Completeness | 62<br>(1.4) | 87<br>(0.9) | + 25<br>(+8, +15)<br>*p<0.001* | 94<br>(1.0) | + 7<br>(-5, 13)<br>*p=0.234* | 95<br>(1.8) | +1<br>(-3, 8)<br>*p=0.324* |
| Error rate | 37<br>(1.7) | 18<br>(1.4) | -17<br>(-25, -13)<br>*p<0.001* | 10<br>(0.7) | -8<br>(-18, -7)<br>*p=0.035* | 8<br>(1.0) | -2<br>(-6, 4)<br>*p=0.180* |

DHIMS2: District Health Information Management System version 2, SD: standard deviation; CI: confidence interval

### Effect on data use for decision making and local planning

At the coaching visit, no health facilities had a facility wall chart or any other means to visualize and use their data for decision making and local planning. However, by the first follow-up visit, 75% of facilities displayed wall charts; this figure rose to 77% at the second follow-up visit (Fig 2). At the first follow-up visit, 84% of new health workers had been oriented to the use of wall charts; this proportion fell slightly to 80% at the second follow-up visit. By the first follow-up visit, 60% of facilities reported using data for decision making and local planning; this rose further to 63% at the second follow-up visit. None of the differences between the first and second follow-up visit were statistically significant.

**Fig 2.**
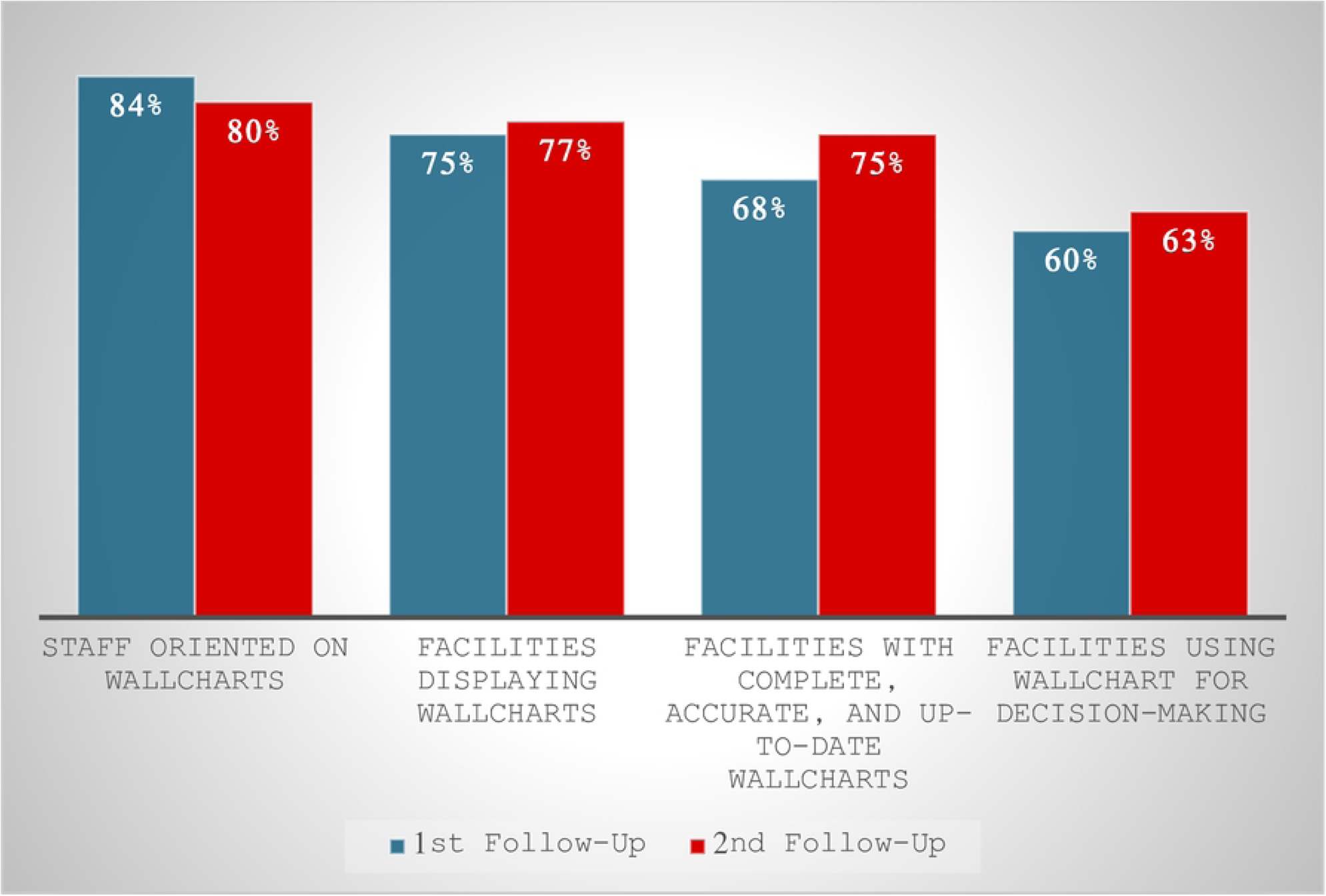
Wall chart availability and use after data coaching (N=231 facilities, N=1211 staff)

### Participant experience survey

A total of 250 health workers, malaria focal persons, and health information officers responded to the qualitative survey to share their experiences with the coaching visit (Table 4). Most were male (73.5%). More than half (63%) of the respondents were aged 30-39 years, with 31% aged 20-29 years and 6% aged 40-49 years. More than half (53%) had between 1 and 5 years of work experience. Their responses reveal how the coaching visit improved data documentation and use by health workers at the CHPS, health centers, district and regional levels.

**Table 4.** Background Characteristics of Participants (N=250)

| Variable | Response(s) | N (%) |
| --- | --- | --- |
| Gender | Female | 66 (26.5) |
|  | Male | 184 (73.5) |
| Age (year(s)) | 20-29 | 78 (31.0) |
|  | 30-39 | 156 (63.0) |
|  | 40-49 | 16 (6.0) |
|  | 50-59 | 0 (0) |
| Years of work experience | <1 | 66 (26.5) |
|  | 1-5 | 133 (53.0) |
|  | 6-10 | 51 (20.5) |

### Improved data quality

Coaching participants roundly agreed that the coaching visits had helped to improve the quality of data documented and reported at their facility. They reported that the coaching had improved their understanding of documentation in the registers, reports, and the data collection process in the facility. Some added that the coaching had helped them to validate their data, and had improved the timeliness of their reporting. Service providers indicated that the wall charts helped them to identify areas for improvement and that being able to see the number of malaria cases suspected, tested and given treatment helped them follow treatment protocols.

> “*The training has helped to improve documentation in the consulting room register and led to timely submission of data.”* - Health information officer

> “*In fact, data coaching visits have increased our knowledge on how to record data properly in the consulting room register, antenatal care register, outpatient register, and reports compilation in our facility.”* - Enrolled Nurse

### Adherence to malaria prevention and treatment guidelines

Respondents reported that wall charts had significantly improved data accuracy, malaria diagnostic accuracy, and staff awareness through promoting improved adherence to guidelines:

> *“Since the implementation of wallc harts to improve adherence to malaria prevention and treatment guidelines, we have observed several improvements and changes among our staff. Here are some notable improvements: Adherence to Test, Treat, Track (T3), reduction of presumptive treatment, improved IPTp and ITN provision, and improved referral practices… Overall, the implementation of wall charts has positively impacted staff adherence to malaria prevention and treatment guidelines. It has promoted evidence-based practices, reduced presumptive treatment, increased adherence to IPTp and ITN provision, and enhanced appropriate referral practices.”* – Senior Technical Officer

> “*Of course, the wall charts have helped facilities adhere to the 3Ts in malaria case management. Facilities can determine the number of malaria cases tested and treated at a glance. And this guides them to monitor their RDT stock levels and put in requisition as soon as possible*.” – Health Information Officer

> *“Since the use of the wall chart, facilities learn to probe further into conditions reported to them and with the appropriate tests conducted. All positive malaria cases are put on antimalarials and asked to come back for review.” -* Nurse

### Local planning

Some service providers reported using wall charts to monitor trends in indicators, both for planning and forecasting. Respondents noted that monitoring trends in indicators gave them advance notice of areas that needed improvement, and that monthly monitoring improves district indicators and identifies errors. Several respondents mentioned that action plans now consider positivity rates and the number of antenatal care registrants in their facilities.

> *“The wall charts help us to identify indicators where we are not meeting the target and to plan…it helps us to monitor trends of health indicators.”* – Health Information Officer

> *“The skills and knowledge I have acquired help me to prepare an action plan for the facility and also help in providing comprehensive care.” –* Registered Nurse

> *“We have improved our district’s indicators due to monitoring on a monthly basis, also enabling us to identify errors with our data before the year ends.”* – Health Information Officer

Respondents frequently mentioned using wall charts to monitor key commodities such as ITNs and antimalarials. Respondents explained that the use of IPTp for guiding commodity allocation has reduced delays in commodity requests and helped monitor commodity stock levels, enabling timely restocking and demand forecasting. Health Information Officers mentioned that learning to monitor stock levels and trends on a monthly rather than annual basis has improved data quality and forecasting for commodities like ITNs and SP, preventing shortages. Wall charts also help facilities notice when commodities are running out, allowing for restocking to avert stockouts:

> *“The coaching has helped us to enter data easily and it’s making our work very simple using the wall chart. At a glance at the graph, I can track our performances and coverage. It makes it easier to do my division and subtraction on the stock using the graph. We are now able to do our requisitions on time so that there will be no shortage of SP for IPTp when pregnant women come for ANC.” -* Registered Nurse

> *“Plans for procurement at the facility now take into consideration the positivity rate recorded whereas ITNs factors in the Number ANC Registrants at a specified period.”* – Health Information Officer

> *“Using the wall chart has been helpful to the facility in monitoring their stock level of commodities … they do not run out of commodities.”-* Health Information Officer

## Discussion

Coaching, supervision, and mentorship approaches have been used to improve health data reporting, analysis, and use in many health areas, but have been rarely detailed in the malaria literature, particularly at the facility level. This study, along with an emerging body of programmatic research, demonstrate that data coaching and other sustained data-focused supportive supervision and mentorship efforts can help improve data literacy, data quality, data visualization, data-driven decision making, and data culture.

The only other documented examples of data coaching for malaria programs come from Madagascar and Uganda. In Madagascar, PMI Measure Malaria developed a data coaching guide that defined the role of a coach, conduct of coaching visits, and performance indicators, and also provided a template to document action items (27). Coaching visits were associated with improved staff skills and confidence using DHIS2, improved data quality (completeness rose from 86 to 91 percent; timeliness rose from 54 to 81 percent), analysis, and visualization. In Kayunga, Uganda, a pilot of an intervention that included in-service training in 5 health centers, mentored continuous improvement with PDSA cycles, learning sessions, and coaching reported improvements in data completeness (28). Health workers appreciated coaching, but some expressed frustration with perceived additional workload from data collection and continuous improvement activities. Financial incentives for participation helped offset this frustration.

Interventions to improve the quality of DHIMS2 data can take technical approaches (e.g., improving paper data collection forms, developing electronic tools, mHealth approaches employing tablets or smartphones, data quality audits/assessments, data storage) or behavioral approaches (e.g., short- or long-term training, task-shifting, supportive supervision, stakeholder engagement, incentives, standardized protocols) (11). Of all interventions, training is the most commonly implemented component, and is widely shown to have a positive impact on data quality. A study of primary health facilities in Nigeria associated training on data management with statistically significant increases in completeness of reporting, accuracy, timeliness, and feedback (29). However, a systematic review showed that interventions with multiple components that include training showed greater benefit than those that tested single intervention components (11). A cross-sectional study of health centers in Chad found that data errors were associated with high workload, stockouts, and unavailability of required data collection tools, as well as the absence of a health technician and dedicated staff for data management (30). These visits also offer opportunities to utilize electronic tools that increase data completeness and reduce errors (31). Across Tanzania, electronic Malaria Services and Data Quality Improvement tools are used during supportive supervision visits to assess the quality of routinely collected malaria data reported from health facilities; the tools include a data quality assessment component, and facilities develop action plans to address deficiencies in service quality identified by the tools (32). Our study showed that data coaching fostered the implementation of functional data management systems in accordance with standards of practice that may strengthen data collection at all levels. Data coaching can also offer an opportunity to evaluate working conditions for clinic personnel, as improvements in workload, staffing, and facility readiness may also improve DHIMS2 data quality.

Strategies that visualize and track health facility indicators—including wall charts, scorecards, and dashboards—are increasingly popular tools to encourage interpretation and use of data for improved performance at the facility and district level. In our study, data coaching activities promoted the creation and display of wall charts, which rose over successive follow-up visits. While studies across health sectors generally demonstrate that data visualization approaches can improve the quality and completeness of data, and improved access to and use of data at district, region, and national levels (21,33,34), their effectiveness in promoting effective use of data at the facility level remains largely unmeasured (35). Most examples are of data being reported upward in the health system, and how data are used is rarely detailed. In Nigeria, a national project to promote use of dashboards using routine data for decision making led to increased completeness of DHIS2 reports from 53% to 81% over three years of the project, but primary healthcare governance structures and limited human resource capacity constrained use of data for decision making (36). In Zanzibar, Jhpiego worked with the Ministry of Health to construct a dashboard integrating multiple sources of malaria-related data (37). The dashboard identified decision-making errors addressed through supportive supervision visits, as well as underutilization of primaquine at private health facilities, prompting the Ministry of Health to develop job aids and antimalarial record books. In Malawi, Hazel et al documented improved reporting consistency and use of data-display wall charts for integrated community case management after a data quality and use package was introduced that included a wall chart template for data display, training for health surveillance assistants and supervisors, and a participatory approach that engaged community and facility health workers in program improvement (21). Some participants reported using data in wall charts for program improvement—for example, to track stock-outs. Sustained support and supervision appear crucial for the success of this modality, as use of the tools declined once scaled up nationally from a smaller pilot. Similarly, use of laminated or electronic wall charts developed in 20 countries under the Maternal and Child Survival Program was sustained in only half of these countries after the project concluded (38). By documenting adoption of wall charts and reports of data use by managers at the second follow-up visit, our study contributes evidence that data coaching visits can empower managers and providers at the facility level, but leadership should consider sustainability of these efforts.

More importantly, surprisingly little research has investigated what data are actually used by managers of primary health care facilities for decisions about service delivery (39). Although the DHIS2 software platform is used to support HMIS in more than 70 countries, including Ghana’s DHIMS2, underutilization or non-use of data by facilities is common (6). Few examples of DHIS2 data use at the local level have been captured in the academic or programmatic literature, particularly for malaria (40). While data can shape effective policy and evidence-based practices, neither possession of nor access to high quality data guarantees its use for decision-making, as a host of organizational factors shape data use (41). For example, in Tanzania, Mboera et al found that while two-thirds of facility respondents used HMIS data, only 38.5% routinely analyzed it due to poor data utilization and inadequate data analysis in most districts and health facilities, stemming from insufficient resources, inadequate incentives and supervision, and nonexistent standard operating procedures for data management (42). A mixed-methods study of prevention of mother-to-child transmission of HIV programs in South Africa found that while 53% of study facilities used data, “use” primarily meant reporting program outputs to the provincial level, not informing decisions and planning (43). The study found an absence of a culture of data use, low trust in the data, and poor data literacy among program and facility managers.

By promoting data culture, which involves designing systems that support work practices and provide forums for conversations around data, data coaching can change health worker perceptions (40). Our findings suggest that data coaching is supporting a culture of data use in Ghana to encourage effective commodity management at facility and district levels, as well as improved case management at the facility level. In Malawi, Kumwenda et al found that health workers providing HIV services saw data management as part of their role, but generally viewed these responsibilities as secondary to provision of clinical care (44). Strategies that motivate health workers to adopt data quality improvement practices at health facilities can promote a data culture that in turn leads to effective data use at the facility level; where workloads are heavy or where private sector providers are involved, financial incentives or recognition can nurture a culture of data use (28,45). Even where health workers articulate the need for an effective health data management system, they may lack technical capacity, policy support, infrastructure, or facility readiness, particularly in rural areas (46). Regular data review meetings, supportive supervision, provision of feedback, and training on computer skills and data collection tools are strategies that can help health workers comprehend the significance and utility of data they collect (33,44).

Sustainable investment in improving data quality requires a systems approach and appropriate governance at all levels of the health system. To make locally appropriate and timely decisions about services, workforce, and procurement, facilities need disaggregated data and managers empowered to create enabling environments (39). Improving the quality of health data requires continuous effort from health facilities, feedback mechanisms between programs, health officers, and providers, and a willingness to make course corrections at all levels (6). Going forward, the impact of data coaching approaches to improve malaria services may be enhanced and made more sustainable through the use of supportive technologies. Electronic tools to improve data quality and data use are increasingly used to supplement training efforts. While many low- and middle-income countries have moved in recent years toward electronic data collection systems to streamline data collection (30), these systems do not ensure overall data quality, nor do they directly promote effective understanding and use of data, particularly at the local level (47). A review of data quality and data use in routine HMIS found that interventions to improve data quality were most effective when they combined capacity building activities such as training, coaching, or supportive supervision with technology enhancement, and that data use for planning rose when interventions promoting data availability were combined with technology enhancement (3). Coupling data coaching with electronic tools for data collection, analysis, and visualization may achieve even greater gains in data quality and data use.

Activities were carried out on a limited budget as part of program activities. We did not evaluate or investigate the organizational culture, infrastructure, or staffing at each study facility, which Daneskohan et al note may have a significant influence on how health workers collect, report, and use data (48). Changes in HMIS error rates and data completeness may be attributable to programmatic or ecological factors other than the data coaching intervention. We relied on self-reports from providers and managers about how data were being used, which may be inflated due to social desirability bias. Additionally, the intervention did not include enhanced technologies that have been shown to magnify the impact of data quality improvement initiatives; this is an area ripe for future research.

## Conclusions

In primary health care facilities in Ghana, a data coaching intervention improved data documentation, management, and reported use for local planning. Participatory development of facility-specific action plans using the PDSA continuous improvement approach promoted local investment in quality improvement. While there are few documented examples of data coaching to improve the quality of malaria surveillance and service data in the academic literature, data coaching—or similar efforts to provide targeted supportive supervision and mentorship on data quality, visualization, and use—offers a model for other malaria intervention programs to document and use HMIS data effectively to improve service quality at the facility level.

## Data Availability

The datasets used and/or analyzed during the current study are available from the corresponding author on reasonable request.

## Acknowledgments

The authors are grateful for the support and expertise of the PMI Impact Malaria in Ghana team. We would especially like to thank the Ghana National Malaria Elimination Program, as well as regional, district, and facility health management teams for their partnership, support, and leadership. The authors would also like to thank Dr. Mark Kabue for his guidance in conceptualizing the manuscript.

## Supporting Information

**S1 Fig. Completed wall chart.**

## Notes

### Competing Interest Statement

The authors have declared no competing interest.

### Funding Statement

This analysis was supported through the U.S. President’s Malaria Initiative (PMI) Impact Malaria project activity (Contract Number 7200AA18C00014) in collaboration with the U.S. Agency for International Development and the U.S. Centers for Disease Control and Prevention. The funders had no role in study design, data collection and analysis, or preparation of the manuscript, but did review and approve the manuscript for submission. The opinions expressed herein are those of the authors and do not necessarily reflect the views of the U.S. President’s Malaria Initiative, the U.S. Agency for International Development, the U.S. Centers for Disease Control and Prevention, or other employing organizations or sources of funding.

## References

1. Mphatswe W, Mate K, Bennett B, Ngidi H, Reddy J, Barker P, et al. Improving public health information: a data quality intervention in KwaZulu-Natal, South Africa. Bull World Health Organ. 2012;90(3).

2. AbouZahr C, Boerma T. Health information systems: the foundations of public health. Bull World Health Organ. 2005;83:578–83.

3. Lemma S, Janson A, Persson LÅ, Wickremasinghe D, Källestål C. Improving quality and use of routine health information system data in low- and middle-income countries: A scoping review. Francis JM, editor. PLoS One [Internet]. 2020 Oct 8 [cited 2023 Mar 18];15(10):e0239683. Available from: https://dx.plos.org/10.1371/journal.pone.0239683

4. Getachew N, Erkalo B, Garedew MG. Data quality and associated factors in the health management information system at health centers in Shashogo district, Hadiya zone, southern Ethiopia, 2021. BMC Med Inform Decis Mak [Internet]. 2022 Dec 1 [cited 2023 Mar 27];22(1):154. Available from: https://bmcmedinformdecismak.biomedcentral.com/articles/10.1186/s12911-022-01898-3

5. Nisingizwe MP, Iyer HS, Gashayija M, Hirschhorn LR, Amoroso C, Wilson R, et al. Toward utilization of data for program management and evaluation: Quality assessment of five years of health management information system data in Rwanda. Glob Health Action. 2014;7(1).

6. Maïga A, Jiwani SS, Mutua MK, Porth TA, Taylor CM, Asiki G, et al. Generating statistics from health facility data: The state of routine health information systems in Eastern and Southern Africa [Internet]. Vol. 4, BMJ Global Health. BMJ Publishing Group; 2019 [cited 2023 Jun 19]. p. 1849. Available from: http://gh.bmj.com/

7. Ohiri K, Ukoha NK, Nwangwu CW, Chima CC, Ogundeji YK, Rone A, et al. An Assessment of Data Availability, Quality, and Use in Malaria Program Decision Making in Nigeria. Health Syst Reform [Internet]. 2016 Oct 1 [cited 2022 Mar 12];2(4):319–30. Available from: http://www.ncbi.nlm.nih.gov/pubmed/31514720

8. Hung YW, Hoxha K, Irwin BR, Law MR, Grépin KA. Using routine health information data for research in low- and middle-income countries: A systematic review [Internet]. Vol. 20, BMC Health Services Research. BioMed Central; 2020 [cited 2023 Mar 28]. p. 1–15. Available from: 10.1186/s12913-020-05660-1

9. Mutale W, Chintu N, Amoroso C, Awoonor-Williams K, Phillips J, Baynes C, et al. Improving health information systems for decision making across five sub-Saharan African countries: Implementation strategies from the African Health Initiative. BMC Health Serv Res [Internet]. 2013 [cited 2023 Mar 30];13(SUPPL.2). Available from: https://pubmed.ncbi.nlm.nih.gov/23819699/

10. Gimbel S, Mwanza M, Nisingizwe MP, Michel C, Hirschhorn L, Hingora A, et al. Improving data quality across 3 sub-Saharan African countries using the Consolidated Framework for Implementation Research (CFIR): Results from the African Health Initiative. BMC Health Serv Res. 2017 Dec 21;17.

11. Lee J, Lynch CA, Hashiguchi LO, Snow RW, Herz ND, Webster J, et al. Interventions to improve district-level routine health data in low-income and middle-income countries: A systematic review [Internet]. Vol. 6, BMJ Global Health. BMJ Publishing Group; 2021 [cited 2023 Jun 19]. Available from: https://pubmed.ncbi.nlm.nih.gov/34117009/

12. Andrada A, Herrera S, Yé Y. Are new national malaria strategic plans informed by the previous ones? A comprehensive assessment of sub-Saharan African countries from 2001 to present. Vol. 18, Malaria Journal. BioMed Central Ltd.; 2019.

13. World Health Organization. Country health information systems: a review of the current situation and trends. Geneva; 2011.

14. World Health Organization. Global technical strategy for malaria 2016-2030, 2021 update [Internet]. Geneva; [cited 2024 Mar 10]. Available from: https://www.who.int/publications/i/item/9789240031357

15. Roll Back Malaria Partnership. Action and Investment to Defeat Malaria 2016-2030 For a Malaria-Free World. Geneva; 2015.

16. World Health Organization. World Malaria Report 2021. Geneva: World Health Organization; 2021.

17. World Health Organization. Analysis and use of health facility data: Guidance for malaria programme managers [Internet]. Geneva; 2018. Available from: https://www.who.int/healthinfo/FacilityAnalysisGuidance_RMNCAH.pdf

18. Ghana Health Service. Ghana Health Service Annual Report. Accra; 2021.

19. Aung T, Niyeha D, Heidkamp R. Leveraging data visualization to improve the use of data for global health decision-making. J Glob Health. 2019;9(2).

20. Karijo EK, Otieno GO, Mogere S. Determinants of Data Use for Decision Making in Health Facilities in Kitui County, Kenya. Quest Journal of Management and Social Sciences. 2021;3(1).

21. Hazel E, Chimbalanga E, Chimuna T, Nsona H, Mtimuni A, Kaludzu E, et al. Using data to improve programs: Assessment of a data quality and use intervention package for integrated community case management in Malawi. Glob Health Sci Pract [Internet]. 2017 Sep 1 [cited 2023 Jun 20];5(3):355–66. Available from: https://pubmed.ncbi.nlm.nih.gov/28963172/

22. Avortri GS, Nabukalu JB, Nabyonga-Orem J. Supportive supervision to improve service delivery in low-income countries: is there a conceptual problem or a strategy problem? BMJ Glob Health [Internet]. 2019 Oct [cited 2023 Mar 27];4(Suppl 9):e001151. Available from: https://pubmed.ncbi.nlm.nih.gov/31673434/

23. Taylor MJ, McNicholas C, Nicolay C, Darzi A, Bell D, Reed JE. Systematic review of the application of the plan-do-study-act method to improve quality in healthcare [Internet]. Vol. 23, BMJ Quality and Safety. BMJ Publishing Group; 2014 [cited 2024 Feb 22]. p. 290–8. Available from: /pmc/articles/PMC3963536/

24. Kobo. KoBoCollect application. Cambridge, MA: KoboToolbox; 2021.

25. StataCorp. Stata Statistical Software: Release 14.2. College Station, TX: StataCorp LP; 2015.

26. ATLAS.ti 7.5 Windows. Berlin, Germany: ATLAS.ti Scientific Software Development GmbH; 2016.

27. PMI Measure Malaria. Coaching and mentorship approach promotes capacity strengthening in reporting, analysis, and use of health data among subnational staff in Madagascar. President’s Malaria Initiative [Internet]. 2021 Apr [cited 2023 Mar 30]; Available from: https://www.measuremalaria.org/news/coaching-and-mentorship-approach-promotes-capacity-strengthening-in-reporting-analysis-and-use-of-health-data-among-subnational-staff-in-madagascar/

28. Hutchinson E, Nayiga S, Nabirye C, Taaka L, Westercamp N, Rowe AK, et al. Opening the ‘black box’ of collaborative improvement: a qualitative evaluation of a pilot intervention to improve quality of malaria surveillance data in public health centres in Uganda. Malar J [Internet]. 2021 Dec 1 [cited 2024 Feb 15];20(1):289. Available from: https://malariajournal.biomedcentral.com/articles/10.1186/s12936-021-03805-z

29. Nwankwo B, Sambo MN. Can training of health care workers improve data management practice in health management information systems: A case study of primary health care facilities in Kaduna State, Nigeria. Pan African Medical Journal [Internet]. 2018 [cited 2023 Jun 19];30. Available from: https://pubmed.ncbi.nlm.nih.gov/30637073/

30. Moukénet A, de Cola MA, Ward C, Beakgoubé H, Baker K, Donovan L, et al. Health management information system (HMIS) data quality and associated factors in Massaguet district, Chad. BMC Med Inform Decis Mak [Internet]. 2021 Dec 1 [cited 2023 Jun 19];21(1). Available from: https://pubmed.ncbi.nlm.nih.gov/34809622/

31. Burnett SM, Wun J, Evance I, Davis KM, Smith G, Lussiana C, et al. Introduction and evaluation of an electronic tool for improved data quality and data use during malaria case management supportive supervision. American Journal of Tropical Medicine and Hygiene [Internet]. 2019 [cited 2023 Jun 19];100(4):889–98. Available from: https://pubmed.ncbi.nlm.nih.gov/30793695/

32. National Malaria Control Programme (NMCP) TanzaniaM of HCDGE and C. Operational manual for implementing malaria services and data quality improvement (MSDQI). Dar es Salaam; 2017.

33. Tchoualeu DD, Elmousaad HE, Osadebe LU, Adegoke OJ, Nnadi C, Haladu SA, et al. Use of a district health information system 2 routine immunization dashboard for immunization program monitoring and decision making, Kano State, Nigeria. Pan Afr Med J [Internet]. 2021 [cited 2023 Jun 19];40(Suppl 1):2. Available from: https://pubmed.ncbi.nlm.nih.gov/36157564/

34. Schulze A, Brand F, Geppert J, Böl GF. Digital dashboards visualizing public health data: a systematic review. Front Public Health [Internet]. 2023 [cited 2023 Jun 20];11:999958. Available from: http://www.ncbi.nlm.nih.gov/pubmed/37213621

35. Buttigieg SC, Pace A, Rathert C. Hospital performance dashboards: a literature review. Journal of Health, Organisation and Management [Internet]. 2017 [cited 2023 Jun 20];31(3):385–406. Available from: https://pubmed.ncbi.nlm.nih.gov/28686130/

36. Etamesor S, Ottih C, Salihu IN, Okpani AI. Data for decision making: using a dashboard to strengthen routine immunisation in Nigeria. BMJ Glob Health [Internet]. 2018 Oct [cited 2023 Jun 19];3(5):e000807. Available from: https://pubmed.ncbi.nlm.nih.gov/30294456/

37. PMI Impact Malaria. Integrated Malaria Dashboard Enhances Data Use for Decision-Making in Zanzibar [Internet]. 2023 [cited 2024 Feb 15]. Available from: https://impactmalaria.org/news-and-blog/posts/integrated-malaria-dashboard-enhances-data-use-for-decision-making-in-zanzibar

38. MCSP. https://mcsprogram.org/across-africa-data-dashboards-in-health-facilities-are-improving-decision-making/. 2019 [cited 2024 Feb 15]. Across Africa, Data Dashboards in Health Facilities are Improving Decision Making. Available from: https://mcsprogram.org/across-africa-data-dashboards-in-health-facilities-are-improving-decision-making/

39. Scott V, Gilson L. Exploring how different modes of governance act across health system levels to influence primary healthcare facility managers’ use of information in decision-making: Experience from Cape Town, South Africa Lucy Gilson. Int J Equity Health [Internet]. 2017 Sep 15 [cited 2023 Jun 19];16(1). Available from: https://pubmed.ncbi.nlm.nih.gov/28911323/

40. Byrne E, Sæbø JI. Routine use of DHIS2 data: a scoping review. BMC Health Serv Res [Internet]. 2022 Dec 1 [cited 2023 Jun 19];22(1). Available from: https://pubmed.ncbi.nlm.nih.gov/36203141/

41. Hoxha K, Hung YW, Irwin BR, Grépin KA. Understanding the challenges associated with the use of data from routine health information systems in low- and middle-income countries: A systematic review [Internet]. Vol. 51, Health Information Management Journal. SAGE Publications Inc.; 2022 [cited 2023 Jun 19]. p. 135–48. Available from: https://pubmed.ncbi.nlm.nih.gov/32602368/

42. Mboera LEG, Rumisha SF, Mbata D, Mremi IR, Lyimo EP, Joachim C. Data utilisation and factors influencing the performance of the health management information system in Tanzania. BMC Health Serv Res [Internet]. 2021 Dec 1 [cited 2023 Jun 19];21(1). Available from: https://pubmed.ncbi.nlm.nih.gov/34030696/

43. Nicol E, Dudley L, Bradshaw D. Assessing the quality of routine data for the prevention of mother-to-child transmission of HIV: An analytical observational study in two health districts with high HIV prevalence in South Africa. Int J Med Inform [Internet]. 2016 Nov [cited 2018 Jul 16];95:60–70. Available from: http://www.ncbi.nlm.nih.gov/pubmed/27697233

44. Kumwenda WWI, Kunyenje G, Gama J, Chinkonde J, Martinson F, Hoffman I, et al. Information management in Malawi’s prevention of mother-to-child transmission (PMTCT) program: Health workers’ perspectives. Malawi Medical Journal [Internet]. 2017 Dec 1 [cited 2023 Jun 19];29(4):306–10. Available from: https://pubmed.ncbi.nlm.nih.gov/29963285/

45. Bennett A, Avanceña ALV, Wegbreit J, Cotter C, Roberts K, Gosling R. Engaging the private sector in malaria surveillance: A review of strategies and recommendations for elimination settings [Internet]. Vol. 16, Malaria Journal. BioMed Central Ltd.; 2017 [cited 2024 Feb 16]. Available from: https://pubmed.ncbi.nlm.nih.gov/28615026/

46. Miiro C, Ndawula JC, Musudo E, Nabuuma OP, Mpaata CN, Nabukenya S, et al. Achieving optimal heath data impact in rural African healthcare settings: measures to barriers in Bukomansimbi District, Central Uganda. Int J Equity Health [Internet]. 2022 Dec 1 [cited 2023 Jun 19];21(1). Available from: https://pubmed.ncbi.nlm.nih.gov/36577986/

47. Kebede M, Adeba E, Adeba E, Chego M. Evaluation of quality and use of health management information system in primary health care units of east Wollega zone, Oromia regional state, Ethiopia. BMC Med Inform Decis Mak [Internet]. 2020 Jun 12 [cited 2023 Jun 19];20(1). Available from: https://pubmed.ncbi.nlm.nih.gov/32532256/

48. Daneshkohan A, Alimoradi M, Ahmadi M, Alipour J. Data quality and data use in primary health care: A case study from Iran. Inform Med Unlocked. 2022 Jan 1;28.

